# Intervention strategies for cystinuria: A systematic review

**DOI:** 10.1101/2020.09.17.20196337

**Authors:** Nirmal P. Bhatt, Aniruddh V. Deshpande, Bernadette Jones-Freeman, Simon H. Jiang, Malcolm R. Starkey

## Abstract

**Purpose:** This systematic review evaluates the current intervention strategies for cystinuria and assesses their quality and efficacy in order to determine the need to identify new and improved strategies for treatment.

**Materials and Methods:** A literature search for interventions in cystinuria was conducted on key electronic databases for studies published between 1996 and 2019. Quality was assessed using Methodological Index for Non-Randomized Studies (MINORS). Studies meeting the inclusion criteria were assessed for study design, patient characteristics and outcomes of interventions, including urinary cystine levels, stone-free rate and stone recurrence rate. A qualitative and critical analysis was performed.

**Results:** Common treatment strategies for cystinuria include hydration and diuresis, alkali therapy and thiol-based therapeutics. Conservative therapies such as adequate hydration and urinary alkalization effectively increased urinary pH, decreased urinary cystine levels and the formation of cystine stones. Second line agents reported included thiols such as Tiopronin, D-penicillamine and captopril. Non-invasive surgical procedures were found to reduce operative trauma and preserved renal function. Combined treatment approaches with hydration and thiols after surgical procedures were associated with less stones and reduced stone recurrence rates. Patient compliance to interventions was often poor and contributed to recurrent cystine stones.

**Conclusions:** Despite existing pharmacological intervention strategies, cystinuria patients frequently require surgical procedures. A more detailed understanding of the mechanisms of pathogenesis of cystinuria as well as an evaluation of patients on an individual basis may be beneficial in reducing the severity of cystinuria, by reducing stone recurrence and associated renal complications.

## INTRODUCTION

Cystinuria is a rare inherited autosomal recessive disease. The estimated overall prevalence of cystinuria is 1 in 7,000 births globally, ranging from 1 in 2,500 in Jewish Israelis and 1 in 100,000 in Sweden^1-3^ Cystinuria accounts for approximately 1-2% of all kidney stone cases in adults and 3-8% in pediatrics.^4-8^ Cystinuria is characterized by defects in the transport of cystine and other dibasic amino acids including lysine, arginine, and ornithine in both the proximal renal tubules and the small intestine. This defect in transportation result in the accumulation of cystine, which precipitates and forms cystine stones.^9^ Cystine stones in the kidney and urinary tract, cause inflammation and obstruction and can predispose an individual to the development of chronic kidney disease.

Cystinuria commonly affects children, and the early age of onset, as well as the high stone recurrence rate, in addition to poor compliance with existing interventions, makes this condition difficult to manage.^10, 11^ Cystinuria is classified into three subtypes denoted type A, type B and type AB. Type A, is caused by mutations in the solute carrier family 3 member 1 (*SLC3A1*) gene encoding the neutral and basic amino acid transport protein (rBAT) heavy subunit.^12, 13^ Type B, is caused by mutations in the solute carrier family 7 member 9 (*SLC7A9*) gene encoding the light subunit b^0,+^ type amino acid transporter 1 (b^0,+^AT).^13, 14^ Type AB occurs when mutations in both *SLC3A1* and *SLC7A9* genes are present.^13^ All three types of cystinuria cause defective reabsorption of cystine and dibasic amino acids by the proximal renal tubule.

Despite the recent developments in the understanding of the genetic causes of cystinuria, ^13, 15, 16^ there is a lack of effective interventions and currently no cure for the disease. Mainstay treatments focus on maintaining cystine homeostasis in the urine. Traditionally, cystinuria management focuses on urinary alkalization using potassium citrate, sodium bicarbonate as well as limiting animal protein intake, whilst maintaining fluid intake to increase urinary volume.^17, 18^ There are currently very limited randomized controlled trials (RCTs) in cystinuria patients that have assessed stone-related outcomes more than 1 year after intervention. Most of the existing studies report combined intervention approaches such as hydration and diuresis and alkali therapy, as well as pharmacological and surgical interventions.

The thiol-based therapeutics alpha-mercaptopropionylglycine (α-MPG or tiopronin), D-penicillamine and captopril are established pharmacological interventions for cystinuria.^17, 19^ Complicated stones may require surgical procedures such as extracorporeal shockwave lithotripsy (ESWL), percutaneous nephrolithotomy (PCNL), ureterorenoscopy (URS), retrograde intrarenal surgery (RIRS) and open surgery (OP) to remove cystine stones.^10, 11, 20^ This review evaluates the existing interventions strategies for cystinuria.

## MATERIALS AND METHODS

### Literature search

A literature search was conducted on key electronic databases including Medline, Embase, Cochrane, Web of Science, Scopus and Google Scholar. English language articles published between 1996 and December 2019 were included. Manual searching of key journals and conference proceedings were performed to retrieve any additional relevant articles. The following search terms were used individually or in combination; cystinuria, cystine stone, cystine calculi, cystine urolithiasis, cystine nephrolithiasis, cystine, kidney, cystine urine and cystinuria patient. The search terms were defined based on the PICO definition; P (population), I (intervention), O (outcomes), while C (comparison) was only applied where placebo-control and internal-control available in the literature.

### Inclusion and exclusion criteria

A diagnosis of cystinuria was based on clinical investigation with at least one previous/current episode of cystine stones, urine cystine levels >250-300mg/L and patients being treated with alkalizing, or pharmacological agents or surgical procedures. The primary outcomes were the type of intervention, change in urinary pH levels, change in urinary cystine levels, stone-free and stone recurrence rates. There were no exclusions based on age, gender, ethnicity, or geographical location. Due to the lack of RCTs for this rare disease, analysis was largely restricted to non-randomized studies with or without control arms. Observational studies including case-control, retrospective and prospective studies were included. Conference abstracts and papers were included if there was sufficient data, and corresponding authors contacted to obtain full study details. Review articles, editorials, news, letters, comments, case series and case reports were excluded. Review articles were used for cross-referencing to retrieve any missing studies and did not contribute to the final number of articles. The inclusion and exclusion criteria were independently applied to all identified articles. Two authors participated in the initial screening of titles and abstracts, and independently screened the titles and abstracts before assessing full text articles. Any disputes were resolved by multiple author agreement. We contacted the authors of the primary reports to request any unclear or unpublished data. If the authors did not reply, the available data was used.

### Data Extraction

This systematic review follows Preferred Reporting Items for Systematic Reviews and Meta-Analysis (PRISMA) guidelines.^21^ References from the database search were pooled into Endnote X8 reference software. Selected data were assessed for study design, aims, inclusion criteria, patient characteristics, intervention strategy and outcomes, renal function data, length of follow-up, stone-free rates, and stone recurrence rates. Intervention studies included alkali therapy, pharmacological agents and surgical procedures outcome comparison at least pre- and post-treatment. The study protocol was registered with PROSPERO to guide this systematic review (ID: CRD42020152061).

### Risk of bias (quality) assessment

Most of the available quality assessment tools are designed for the evaluation of RCTs, case-control or cohort studies. Here, the methodological quality assessment tool, Methodological Index for Non-Randomized Studies (MINORS), was used to assess the studies that met the inclusion criteria. MINORS is validated tool applied for the methodological quality of non-comparative and comparative observational studies.^22^ It consists of 8 items for non-comparative and an additional 4 items for comparative studies. The quality assessment were performed independently by 2 authors (NPB and MRS). AVD, BJF and SHJ cross-checked the selected literature and quality assessment tool to resolve any disputes.

### Data synthesis

Due to the anticipated variability in the included studies, and lack of RCTs, meta-analysis was not planned. Hence a qualitative and critical analysis of the data was performed with available patient characteristics, disease history, and treatment procedures and effectiveness. Post-treatment outcomes as mentioned above were compared with patient’s history.

## RESULTS

### Search results

The initial database and Google search retrieved a total of 1445 articles. After excluding duplicate studies, 660 studies were reviewed for title and abstract screening. 594 were excluded due to being non-relevant (500), a review (34), case reports (48), animal studies (4), supplementary comments (3), non-English (2) or published prior to 1996 (3). A total of 66 studies underwent full-text review. 40 articles were excluded after screening the full text; non-relevant studies (20), inappropriate study aims or outcomes (15) or full text not available (5). 24 studies met the review criteria and the MINORS quality assessment tool was applied to these studies, which resulted in the exclusion of 2 studies that did not meet the quality control requirements (Figure 1).

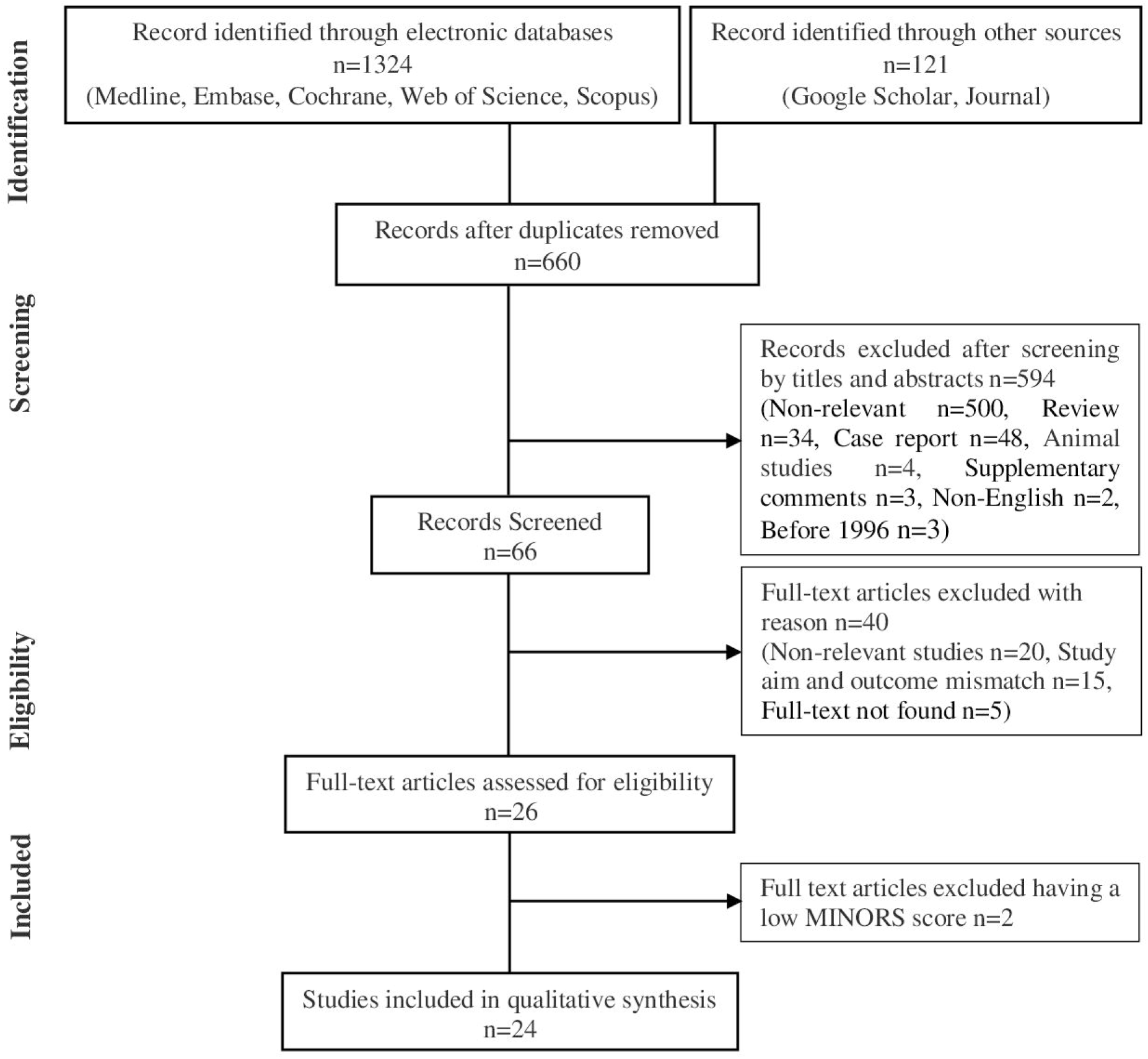

### Study characteristics

A summary of the study characteristics is provided in Table 1. 17 studies were retrospective observational studies and 7 were prospective observational studies, with or without comparison groups. Of the 24 total studies, 11 were non-comparative studies and 13 were comparative studies. The quality of the included studies were assessed using the MINORS score (Table 2 and 3). The mean score for non-comparative studies was 11 out of 16 (range, 8-13). For non-comparative studies MINORS scores are classified as: low (score 5-8), moderate (score 9-12), and high quality (score 13-16). The mean score for comparative studies was 17 out of 24 (range, 12-21). For comparative studies MINORS scores are classified as: low (score 7-12), moderate (score 13-18) and high quality (score 19-24). Those studies that scored a moderate or high MINORS score were included. 2 studies were excluded due to a low MINORS score.

**Table 1.** Summary of the study characteristics.

| **Study reference (alphabetical) and study type** | **Number of patients (male (M) and Female (F)) and mean or median age (years)** | **Interventions (%)** | **Follow-up period (years)** | **Total number of stone episodes and/or surgical interventions** | **Stone episodes per patient and/or stones per patient-year and/or stone recurrence per patient-year** | **Total surgical procedures per patient and/or per patient-year** | **Stone-free after treatment (%)** | **Therapy successful (Yes/Partially)** |
| --- | --- | --- | --- | --- | --- | --- | --- | --- |
| Ahmed *et al.* 2006<br>Retrospective observational | 30 (16M and 14F) cystinuria patients<br>Mean age = 39 | 21 (80.7%) Potassium citrates, 17 (65.3%) Sodium bicarbonate, 11 (42.3%) D-penicillamine<br>143 (61%) ESWL, 28 (12%) PCNL, 50 (21%) URS and 16 (6%) OP | Mean = 6 | 126 stone episodes and 237 surgical procedures | 4.2 stone episodes per patient | 7.9 per patient | 28/30 (93.3%) | Yes |
| Akakura *et al.* 1999<br>Retrospective observational | 31 (22M and 9F) cystinuria patients<br>Mean age = 33 | Potassium/Sodium citrate, Sodium bicarbonate, 28 (90.3%) D-penicillamine and Tiopronin, 2 (6.5%) Captopril | Mean = 7.5 | N/A | 0.19 stones per patient-year | N/A | 15 (48.4%) | Yes |
| Assimos *et al.* 2002<br>Prospective observational | 85 (49M and 36F) cystinuria patients vs 3,964 (2,594M | ESWL, PCNL, OP | N/A | N/A | N/A | 14.1% (cystinuria patients) and 2.96% (CaOx stone patients) procedures, | N/A | Yes |
| | and 1,370F)<br>CaOx stone patients<br>Cystinuria patients<br>mean age = 42.6<br>CaOx stone patients<br>mean age = 48.9 | | | | | $p=0.007$ | | |
| Barbey *et al.* 2000<br>Retrospective observational | 27 (12M and 15F)<br>cystinuria patients<br>Mean age = 19.6 | 5 (18.5%) Potassium citrate, 22 (81.4%) Sodium bicarbonate, (8 - 18gm/kg/day)<br>D-penicillamine (600-1200mg/day),<br>Tiopronin (500-1000mg/day),<br>Captopril (100-150mg/day)<br>32 (25.6%) SWL, 15 (12%) PCNL, 7 (5.6%) RIRS and 65 (53.3%) OP | Mean = 11.6 | 66 stone episodes and 44 surgical procedures | 2.4 stone episodes per patient and 0.20 stones per patient-year | 1.5 per patient and 0.14 per patient-year | 13 (48%) | Yes |
| Chow *et al.* 1996<br>Retrospective observational | 16 (9M and 7F)<br>cystinuria patients<br>Mean age = 33.1 | Potassium citrate, Sodium bicarbonate, D-penicillamine (1000-2000gm/day) or Tiopronin (800-1,200mg/day) in divided doses, | Mean = 6.5 | 88 stone episodes and 34 surgical procedures | 5.5 stone episodes per patient and 0.84 stones per patient-year | 2.1 per patient and 0.3 per patient-year | 12/16 (75%) | Yes |
|  |  | Captopril (50mg 3 times/day)<br>10.5% ESWL,<br>68.4% PNCL, 21% RIRS |  |  |  |  |  |  |
| Chow *et al.* 1998<br>Retrospective observational | 31 (14M and 17F) cystinuria patients<br>Mean age = 33.8 | Potassium citrate, Sodium bicarbonate, D-penicillamine (1000-2000gm/day) or Tiopronin (800-1,200 mg/day), Captopril (50mg 3 times day)<br>9 (14.8%) ESWL (stone <1.5 cm<sup>2</sup>), 41 (67.2%) PCNL (stone > 1.5 cm<sup>2</sup>), 7 (11.5%) RIRS, 4 (6.6%) PCNL+ESWL | Mean = 4.6 | 61 surgical procedures | N/A | 1.98 per patient | 52 (86.90%) | Yes |
| DeBerardinis *et al.* 2008<br>Retrospective observational | 11 cystinuria patients<br>Mean age = 7.4 | D-penicillamine (5-20 mg/kg/day) | Mean = 9 | N/A | N/A | N/A | N/A | Partially |
| Fjellstedt *et al.* 2001<br>Prospective observational | 14 (8M and 6F) cystinuria patients<br>Mean age = 21 | 13 Potassium citrate (60mmol/day), Sodium bicarbonate (71.4mmol/day), 10 Tiopronin (1250-3000mg/day) 7 ESWL | Mean = 0.1 | N/A | N/A | N/A | N/A | Yes |
| Izol *et al.* 2013<br>Prospective case-control | 17 (10M and 7F) cystinuria patients<br>Mean age = 8.5 | Potassium citrate and Citric acid<br>SWL, PCNL | Mean = 2.5 | N/A | N/A | N/A | N/A | Yes |
| Malieckal *et al.* 2019<br>Prospective observational | 10 (7M and 3F) cystinuria patients<br>Mean age = 49 | 3 (30%) D-penicillamine, 7 (70%) Tiopronin | N/A | N/A | N/A | N/A | N/A | Yes |
| Moore *et al.* 2018<br>Retrospective observational | 16 (8M and 8F) cystinuria patients<br>Median age = 15.5 | Potassium citrate<br>D-penicillamine (500mg twice a day)<br>PCNL, SWL, URS | Mean = 8.6 | N/A | 62.5% of patients had recurrent stones | 3.1 per patient | N/A | Yes |
| Onal *et al.* 2013<br>Retrospective observational | 51 (29M and 22F) cystinuria patients<br>Median age = 6 | Potassium citrate or Potassium sodium hydrogen citrate (1mEq/kg),<br>Tiopronin (10mg/kg)<br>11 (16.9%) SWL, 14 (21.5%) PCNL, 2 (3.1%) URS, 4 (6.1%) OP, 1 (1.5%) PCNL+OP | Median = 8 | N/A | N/A | N/A | 73.8% | Yes |
| Pareek *et al.* 2005<br>Retrospective observational | 20 (6M and 14F) cystinuria patients<br>Mean age = | 5 vs 16 PCNL, 5 vs 15 Ureteroscopy, 1 vs 5 SWL in complaint and non-complaint patients | Mean = 3.5 | N/A | N/A | 1 (complaint) and 4 (non-complaint) per patient | 8 (73%) complaint and 3 (33%) non- | Yes |
|  | 45 |  |  |  |  |  | complaint |  |
| Pietrow *et al.* 2003<br>Retrospective observational | 26 (11M and 15F) cystinuria patients<br>Mean age = 32 | Tiopronin (1000mg/day) | Mean = 3.2 | N/A | N/A | N/A | N/A | Partially |
| Shim *et al.* 2014<br>Retrospective observational | 14 (8M and 6F) cystinuria patients<br>Mean age = 19.6 | 9 (64.3%) Potassium citrate (1000 mg 3 times/day), Sodium bicarbonate (500mg 4 times/day), 3 (21.4%) D-penicillamine (250mg twice a day)<br>12 PNCL, 25 URS, 1 OP | Median = 5 | 38 surgical procedures | 2.7 stone episodes per patient and 3.2 stone recurrences per patient-year | N/A | 9 (64.2%) | Yes |
| Sfoungaristos *et al.* 2015<br>Retrospective case-control | 30 (18M and 12F) cystinuria patients<br>Cystinuria patients mean age = 45.6<br>CaOx stone patients mean age = 46 | PCNL, ESWL, URS, RIRS | N/A | 9.80±9.17 stone episodes for cystinuria patients and 2.73±1.95 for CaOx stone patients | N/A | N/A | N/A | Yes |
| Strologo *et al.* 2007<br>Prospective observational | 20 cystinuria patients<br>Mean age = 12.6 | Potassium citrate, Sodium bicarbonate, Tiopronin (25 mg/kg/day), D-penicillamine (17mg/kg/day)<br>PCNL, OP | Mean = 3.5 | N/A | 0.28 stones per patient-year | N/A | N/A | Yes |
| Tekin *et al.* 2001<br>Prospective case-control | 18 (13M and 5F) cystinuria patients vs 24 (17M and 7F) healthy controls<br>Median age = 6.5 | Potassium citrate (1mEq/kg/day), Tiopronin (10-15mg/kg/day)<br>2 (11.1%) ESWL, 2 (11.1%) PCNL, 9 (50%) OP, 3 (16.6%) ESWL+OP, 2 (11.1%) ESWL+PCNL, 1 (5.55) URS | Median = 1.3 | N/A | 0.64 stones recurred per patient-year | N/A | 12/18 (66.7%) | Yes |
| Trinchieri *et al.* 2007<br>Retrospective observational | Traditional cystinuria treatment group 31 (18M and 13F) vs less invasive cystinuria treatment group 17 (8M and 9F)<br>Mean age = 37.2 | PNCL, ESWL, OP | N/A | 20.3 stone episodes in traditional and 12.1 stone episodes in less invasive treatment group | Stone recurrence per patient-year was 1.34 in traditional and 1.16 in less invasive surgical group | N/A | N/A | Yes |
| Usawachintachit *et al.* 2018<br>Retrospective case-control | 42 (20M and 22F) cystinuria patients<br>Median age = 18.5 | 36 (85.7%) Potassium citrate<br>11 (26.2%) Tiopronin, 2 (4.8%) D-penicillamine | Mean = 8.8 | 261 surgical procedures | N/A | 5 per patient and 0.57 per patient-year | N/A | Yes |
| Varda *et al.* 2015<br>Retrospective observational | Median age = 12 | 13 PCNL, 44 URS, 16 ESWL, 3 OP | Median = 4.5 | 110 surgical procedures | 6 stone episodes per patient and 1 stones per patient-year | N/A | N/A | Yes |
| Worcester *et al.* 2006<br>Prospective case-control | 52 (24M and 28F) cystinuria patients vs 3215 non-cystine stone patients<br>Cystinuria patients mean age = 34<br>Non-cystine stone patients mean age = 43 | Potassium citrate 9 D-penicillamine or Captopril, 20 Tiopronin<br>SWL | Mean = 4.3 | N/A | N/A | 3.2-4.8 (cystinuria patients) vs 0.60-1.16 (CaOx stone patients) per patient | N/A | Yes |
| Yamacake *et al.* 2016<br>Retrospective observational | 8 (2M and 6F) cystinuria patients<br>8 (2M and 6F) struvite stone patients<br>Mean age =<br>24.7 | Cystine stone: 7/8 PCNL, SWL and OP<br>Struvite stone: 5/8 PCNL, SWL and OP | Cystinuria patients mean = 4.7<br>Struvite stone patients mean = 3.3 | N/A | N/A | 3.0 (cystinuria) and 2.2 (struvite stone patient) per patient | 70% (cystine stone) and 80% (struvite stone) | Yes |
| Yuruk *et al.*<br>2017<br>Retrospective<br>observational | 14<br>cystinuria<br>patients<br>Mean age =<br>10.9 | Potassium citrate,<br>Tiopronin<br>RIRS | Mean = 2.1 | N/A | N/A | N/A | 100% | Yes |

SWL/ESWL, extracorporeal shockwave lithotripsy; PCNL, percutaneous nephrolithotomy; RIRS, retrograde intrarenal surgery; URS, ureterorenoscopy; OP, open surgery; CaOx, calcium oxalate; N/A, not available

**Table 2.** Quality assessment for non-comparative studies.

| Study<br>Items | Ahmed *et al.*<br>2006 | Barbey *et al.*<br>2000 | Chow *et al.*<br>1996 | Chow *et al.*<br>1998 | DeBerardinis<br>*et al.* 2008 | Onal *et al.*<br>2013 | Shim *et al.*<br>2014 | Strologo *et al.*<br>2007 | Verda *et al.*<br>2015 | Worcester *et al.* 2006 | Yuruk *et al.*<br>2017 |
| --- | --- | --- | --- | --- | --- | --- | --- | --- | --- | --- | --- |
| **A clearly stated aim** | 2 | 2 | 2 | 2 | 2 | 2 | 2 | 2 | 2 | 2 | 2 |
| **Inclusion of consecutive patients** | 2 | 2 | 2 | 2 | 2 | 2 | 2 | 2 | 2 | 2 | 2 |
| **Prospective collection of data** | 0 | 2 | 1 | 1 | 0 | 1 | 1 | 1 | 1 | 1 | 2 |
| **Endpoints appropriate to the aim of the study** | 2 | 2 | 1 | 2 | 2 | 1 | 2 | 2 | 1 | 2 | 2 |
| **Unbiased assessment of the study endpoint** | 1 | 1 | 1 | 1 | 1 | 1 | 1 | 1 | 1 | 2 | 1 |
| **Follow-up period appropriate to the aim of the study** | 2 | 2 | 2 | 2 | 2 | 2 | 2 | 2 | 2 | 2 | 2 |
| **Loss of follow-up less than 5%** | 2 | 1 | 2 | 2 | 1 | 1 | 1 | 1 | 2 | 2 | 2 |
| **Prospective calculation of the study size** | 0 | 0 | 0 | 0 | 0 | 0 | 0 | 0 | 0 | 0 | 0 |
| **TOTAL SCORE (/16)** | 11 | 12 | 11 | 12 | 10 | 10 | 11 | 11 | 11 | 13 | 13 |
| **Study quality** | M | M | M | M | M | M | M | M | M | H | H |

H, High; M, Moderate; L, Low

**Table 3.** Quality assessment for comparative studies.

| **Study**<br>**Items** | **Akakura**<br>***et al.***<br>**1999** | **Assimos**<br>***et al.***<br>**2002** | **Fjellstedt**<br>***et al.***<br>**2001** | **Izol *et al.***<br>**2013** | **Malieckal**<br>***et al.***<br>**2019** | **Moore *et al.***<br>**2018** | **Pareek *et al.***<br>**2005** | **Pietrow *et al.***<br>**2003** | **Sfougari**<br>**stos *et al.***<br>**2015** | **Tekin *et al.***<br>**2001** | **Trinchieri**<br>***et al.***<br>**2007** | **Usawachi**<br>**ntachit *et al.***<br>**2018** | **Yamacak**<br>***et al.***<br>**2016** |
| --- | --- | --- | --- | --- | --- | --- | --- | --- | --- | --- | --- | --- | --- |
| **A clearly stated aim** | 2 | 2 | 2 | 2 | 2 | 2 | 2 | 2 | 2 | 2 | 2 | 2 | 2 |
| **Inclusion of consecutive patients** | 2 | 2 | 2 | 2 | 2 | 2 | 2 | 2 | 2 | 2 | 2 | 2 | 2 |
| **Prospective collection of data** | 1 | 1 | 1 | 1 | 2 | 1 | 2 | 0 | 0 | 1 | 1 | 2 | 2 |
| **Endpoints appropriate to the aim of the study** | 2 | 2 | 1 | 2 | 2 | 2 | 2 | 2 | 2 | 2 | 2 | 2 | 2 |
| **Unbiased assessment of the study endpoint** | 2 | 1 | 2 | 1 | 2 | 2 | 2 | 1 | 2 | 2 | 2 | 1 | 1 |
| **Follow-up period appropriate to the aim of the study** | 2 | 0 | 1 | 2 | 0 | 2 | 2 | 2 | 0 | 1 | 0 | 2 | 1 |
| **Loss of follow-up less than 5%** | 2 | 0 | 1 | 2 | 0 | 2 | 2 | 1 | 2 | 2 | 0 | 2 | 2 |
| **Prospective calculation of the study size** | 0 | 0 | 0 | 0 | 0 | 0 | 0 | 0 | 1 | 0 | 0 | 0 | 0 |
| **An adequate control group** | 0 | 2 | 2 | 1 | 0 | 0 | 1 | 0 | 2 | 2 | 1 | 2 | 2 |
| **Contemporary groups** | 2 | 2 | 1 | 2 | 2 | 2 | 2 | 1 | 2 | 2 | 1 | 2 | 2 |
| **Baseline** | 1 | 1 | 1 | 1 | 2 | 2 | 2 | 2 | 2 | 1 | 1 | 2 | 2 |
| **equivalence of groups** |  |  |  |  |  |  |  |  |  |  |  |  |  |
| **Adequate statistical analysis** | 1 | 2 | 2 | 1 | 1 | 2 | 2 | 2 | 2 | 1 | 2 | 2 | 1 |
| **TOTAL SCORE (/24)** | 17 | 15 | 16 | 17 | 15 | 19 | 21 | 15 | 19 | 18 | 14 | 21 | 19 |
| **Study quality** | M | M | M | M | M | H | H | M | H | M | M | H | H |

H, High; M, Moderate; L, Low

Of the 24 studies, 18 reported outcomes from alkalinizing compounds (potassium citrate and sodium bicarbonate) and/or thiol drugs (D-penicillamine, tiopronin and captopril).^10, 11, 17-20, 23-34^ 20 studies reported outcomes from surgical interventions in cystinuria patients.^10, 11, 17-20, 24-26, 29, 30, 32-40^ Of the 24 studies, 15 reported combined intervention methods with alkalinizing compounds, pharmacological drugs and/or surgical procedures.^10, 11, 17-20, 24-27, 29, 30, 32-34^ No studies directly compared pharmacological intervention to surgical intervention.

In this review, a total of 654 cystinuria patients (mean 27.25 per study) were included. Among these patients, 323 were male (53%) and 286 (47%) were female, from 21 studies. 3 studies did not distinguish patient’s gender. 19 studies reported mean patient age, which ranged from 7.4 to 49 years.^10, 17-20, 23-28, 31, 32, 34-38, 40^ Of the 24 studies, 5 reported median patient age, which ranged from 6 to 18.5 years.^11, 29, 30, 33, 39^

### Effectiveness of urine dilution and alkalization

Initial treatment for cystinuria is to maintain daily urine volume >3L/day.^17^ In addition to appropriate hydration, urinary alkalization is used to maintain urinary pH >6.5.^18^ Of the 24 studies, 16 reported the use of potassium citrate and sodium bicarbonate as an alkalizing agents for the management of cystinuria.^10, 11, 17-20, 24-27, 29-34^ 5 studies reported the detail concentration of alkalizing agents used and their outcome of treatment (Table 4).^11, 17, 18, 26, 32^ Potassium citrate (1mEq/kg/day,^11, 30^ 40-70mmol/day,^18^ 1000mg three times/day^32^) and sodium bicarbonate (8-18gm/kg/day,^17^ 47.6-107mmol/day,^18^ 500mg four times/day^32^) were recommended for all patients after cystinuria diagnosis. A significant increase in urinary pH was observed with sodium bicarbonate treatment (6.4-7.25, *p*<0.01),^18^ and potassium citrate treatment (6.4-7.10, *p*<0.05),^18^ (5.6-6.9, *p*=0.02)^11^ and (5.8-7.55, *p*<0.001).^26^ Post-treatment urinary pH levels were increased >7 with treatment of potassium citrate and sodium bicarbonate.^17, 32^

**Table 4.** Effectiveness of urine dilution and alkalization.

| Study reference | Interventions | pH change | Stone recurrence % | Key outcomes |
| --- | --- | --- | --- | --- |
| Barbey *et al.* 2006 | Potassium citrate and sodium bicarbonate (8-18gm/kg/day) | 7.5 | 78 | Increased urinary pH and reduced urinary cystine levels |
| Fjellstedt *et al.* 2001 | Potassium citrate and sodium bicarbonate (40-70mmol/day and 47.7-107mmol/day) | 6.4-7.10, $p<0.05$ (potassium citrate) and 6.4-7.25, $p<0.01$ (sodium bicarbonate) | NR | In the absence of severe renal impairment, alkalization significantly increases urinary pH levels |
| Izol *et al.* 2013 | Potassium citrate and citric acid (2mmol/kg per day in 3 divided doses) | $5.8\pm0.5$-$7.5\pm0.4$, $p<0.001$ | 41 | Increased urinary pH and reduced stone recurrence rate |
| Shim *et al.* 2014 | Potassium citrate and sodium bicarbonate (1000mg 3 times/day and 500mg 4 times/day) | 6.5-7.2 | NR | Increased urinary pH levels |
| Tekin *et al.* 2014 | Potassium citrate (1mEq/kg/day) | $5.6\pm0.2$-$6.9\pm0.3$, $p=0.020$ | 33 | Increased urinary pH and decreased urinary citrate excretion |

Cystinuria patients excrete less citrate (262±428mg/1.73/m3) than healthy controls (491±490mg/1.73/m3, *p*=0.044) with urinary citrate excretion in the pre-treatment group lower (255±219mg./1.73/m3) than the post-treatment group (729±494mg./1.73/m3, *p*=0.003).^11^ Urinary citrate excretion was also increased by alkalization therapy (2.9±1.1-5.4±1.3mmol/day, *p*<0.01).^17^

Urinary sodium excretion was significantly increased (144mmol/day-220mmol/day, *p*<0.05)^18^ and (173±58mmol/day-263±91mmol/day, *p*<0.001)^17^ by sodium bicarbonate treatment. Urinary potassium excretion (63mmol/day-94mmol/day, *p*<0.01) were also significantly increased during treatment with potassium citrate.^18^

### Effectiveness of pharmacological interventions

17 studies reported pharmacological intervention using the thiol-based drugs; tiopronin, D-penicillamine and captopril.^10, 11, 17-20, 23-25, 27-34^ These interventions were applied when hydration and alkalization treatment was not effective in reducing urinary pH and cystine levels. Of the 17 studies, 12 reported urinary pH change, urinary cystine levels, number of stones per patient-year and stone free rate (Table 5).^11, 17, 19, 23-25, 27-32^

**Table 5.** Effectiveness of pharmacological interventions.

| Study reference | Interventions | pH change | Urinary cystine levels | No. of stones per patient-year | Stone-free % | Key outcomes |
| --- | --- | --- | --- | --- | --- | --- |
| Akakura *et al.* 1999 | Tiopronin, D-penicillamine, captopril | NR | 221.2±75.2mg/L | 0.19 | 48 | Decreased urinary cystine levels and lower stone events per year |
| Barbey *et al.* 2000 | Tiopronin (500-1000mg/day), D-penicillamine (600-1200mg/day), captopril (100-150mg/day) | 7.5 | No thiols<br>808±305mg/24 hours<br>and 585±128mg/24 hours with thiols,<br>$p=0.046$ | 0.20 | 22 | Decreased urinary cystine levels and stone recurrence rate |
| Chow *et al.* 1996 | Tiopronin (800 to 1,200mg/day, D-penicillamine (1000-2000gm/day), captopril (50 mg 3 times/day) | >7.5 | 771mg/24 hours | 0.84 | 75 | 65% of cystinuria patients had decreased stone formation |
| Chow *et al.* 1998 | Tiopronin (800-1,200 mg/day), D-penicillamine (1000-2000gm/day), captopril (50 mg 3 times day) | NR | NR | NR | 87 | Decreased stone recurrence rate when urinary cystine excretion <797 mg/gm creatinine/24 hours |
| DeBerardinis *et al.* 2008 | D-penicillamine (5-20 mg/kg/day) | NR | Decreased by 54% | NR | NR | Decreased urinary cystine levels |
| Malieckal *et al.* 2019 | Tiopronin, D-penicillamine | NS | Reduced 24 hours urinary cystine excretion, $p<0.039$ | NR | NR | Decreased urinary cystine levels |
| Moore *et al.* 2018 | D-penicillamine (500mg twice a day) | NR | NR | NR | 63 | Successful stone clearance was associated with decreased stone recurrence rate |
| Onal *et al.* 2013 | Tiopronin (10 mg/kg/day) | NR | NR | NR | 74 | Tiopronin was recommended for cystine stone patients after stone analysis |
| Pietrow *et al.* 2003 | Tiopronin (1000mg/day) | NR | 154.3mg/L, $p=0.004$ | 0.5 | NR | Decreased urinary cystine levels |
| Shim *et al.* 2014 | D-penicillamine (250 mg twice a day) | NR | NR | 3.2 during 5 years follow-up | 64 | 64.3% of cystinuria patients were compliant with medical treatment resulting in significantly reduced residual stones |
| Strologo *et al.* 2007 | Tiopronin (25 mg/kg/day), D-penicillamine (17mg/kg/day) | NR | NR | Stone events decreased from 0.28 to 0.03 | NR | Decreased stone events per patient |
| Tekin *et al.* 2001 | Tiopronin (10-15 mg/kg/day) | 5.6±0.2-6.9±0.3, $p=0.020$ | 245±266 mmol/mol.creatinine and 140±106 mmol/mol.creatinine, $p=0.015$ | NR | 67 | Decreased urinary cystine levels |

7 studies reported an association between pharmacological interventions and urinary cystine excretion, along with additional outcomes such as the number of stone episodes, the number of surgical procedures, stone recurrence rate and stone-free rate.^11, 17, 23, 24, 27, 28, 31^ Urinary cystine decreased from 1,052±161mg/day to 755±81mg/day with tiopronin treatment, and from 789±126mg/day to 517±92mg/day with D-penicillamine in the mean patients age of 19.6 years. However, captopril did not change cystine excretion (1,044±57mg/day to 1,039±137mg/day).^17^ A significant difference was reported between patients treated with hyperdiuresishyper-diuresis and alkalization, and added thiols (808±305mg/24hrs *vs* 585±128mg/24hrs, *p*=0.046).^17^ The average urinary cystine levels were reduced by 54% (range, 5-81%) with D-penicillamine (20mg/kg/day),^23^ and 15% of cystinuria patients maintained urinary cystine excretion <300mg/L following tiopronin (1000mg/day) treatment.^31^ Further, two studies reported a significant reduction in urinary cystine levels (*p*=0.015)^11^ and (*p*<0.039)^28^ following intervention with tiopronin and/or D-penicillamine. Decreased urine cystine level (221.2±75.2mg/L) was reported when stone events <0.3 per year compared to increased urine cystine level (303.3±93.5mg/L) when stone events ≥0.3 per year).^27^ Cystinuria patients had less urinary citrate excretion compared to the control group (262±428mg/1.73/m3-491±490mg/1.73/m3, *p*=0.044), however, citrate excretion was increased (*p*=0.003) after intervention with tiopronin (10-15mg/kg/day).^11^

Cystinuria patients with alkalization and thiol intervention had up to a 65% reduction in yearly stone events, which equated to 2.7 per patient or 0.52 stone events per patient-year.^19^ Tiopronin (13.8-51mg/kg/day) and D-penicillamine (16.3-17.8mg/kg/day) reduced stone events from 0.28 to 0.03 per year.^24^ Stone free rates ranged from 22 to 87% with diuresis, alkalization and pharmacological management after single or multiple surgical procedures.^11, 17, 19, 25, 27, 29, 30, 32^ Thiol-treated patients had decreased stone recurrence when urinary cystine excretion was maintained at <797 mg/gm creatinine/24 hours.^25^

### Effectiveness of surgical interventions

Surgical intervention strategies for cystine stones include ESWL, PCNL, URS, RIRS and OP. Either single or combined surgical procedures were partially effective in removing stones.^10, 11, 17-20, 24-26, 29, 30, 32-40^ Ahmed *et al*. reported cystine stone patients needed repeated surgical procedures to remove stones including ESWL which accounted for 61%, URS 21%, PCNL 12% and OP 6% of surgical procedures.^10^ Another study reported ESWL 26%, PCNL 12%, RIRS 6% and OP 53% surgical procedures were required to remove cystine stones.^17^ Typically, males required more surgical procedures than females (2.9 *vs* 2.5).^32^

Of the 20 studies, 13 reported detail about intervention type, stone position, intervention procedure, stone free rate and stone recurrence rate (Table 6).^10, 11, 17, 19, 20, 25, 29, 30, 32, 33, 36, 38, 39^ The type of surgical treatment selected was dependent on the size and location of the stones. 5 studies reported various stone positions from unilateral, bilateral, left and right kidney, calyceal, pelvic and ureteral cystine stones.^17, 19, 30, 32, 33^ Bilateral cystine stones were associated with a higher number of lifetime stone surgeries than unilateral stones (6.5 *vs* 2 sessions, *p*<0.05).^33^ Stones were found in the left (46%) and right (54%) kidney.^30^ Furthermore, 41% calyceal, 55% ureter and 4% pelvic stones were present in patients with cystinuria.^32^

**Table 6.**
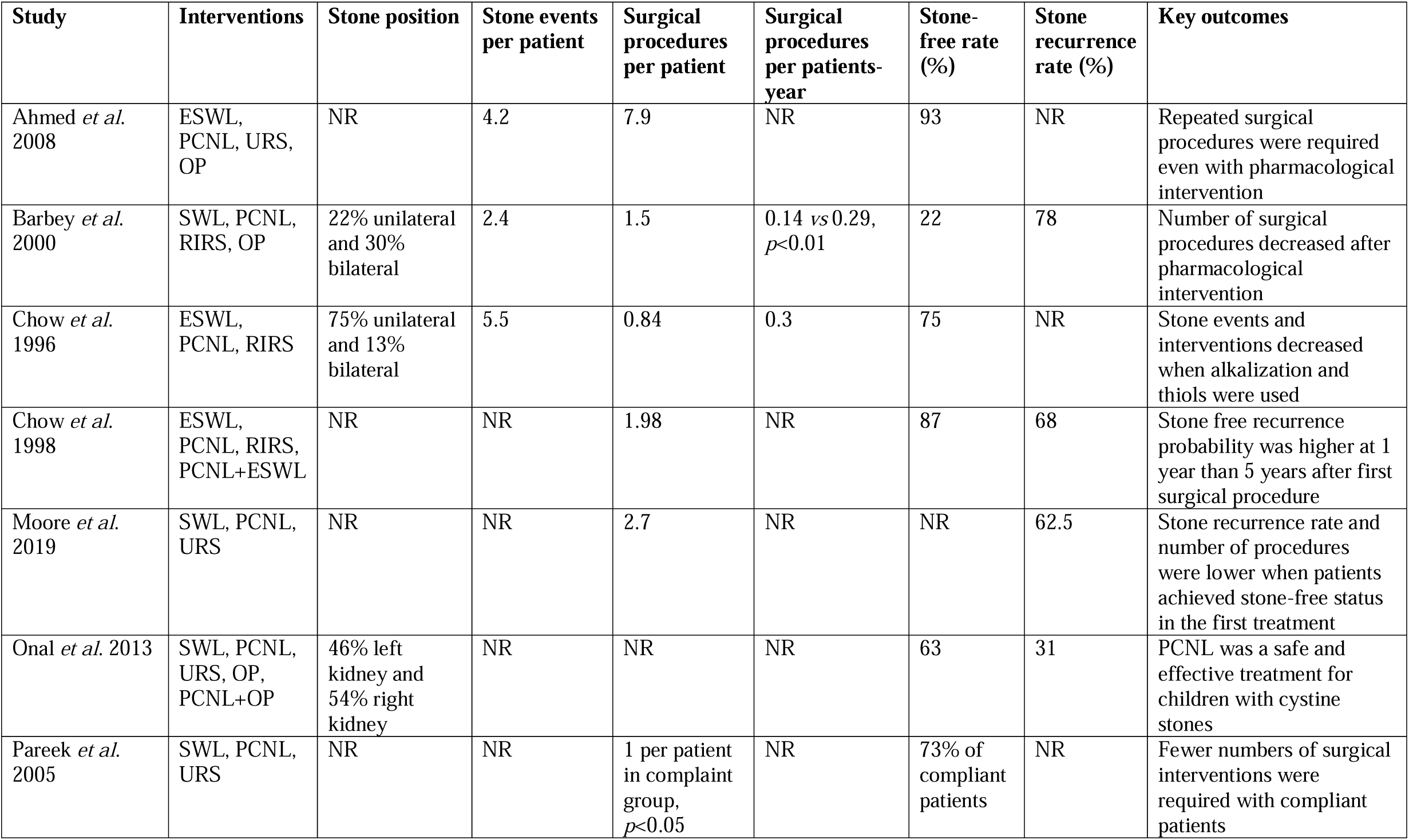
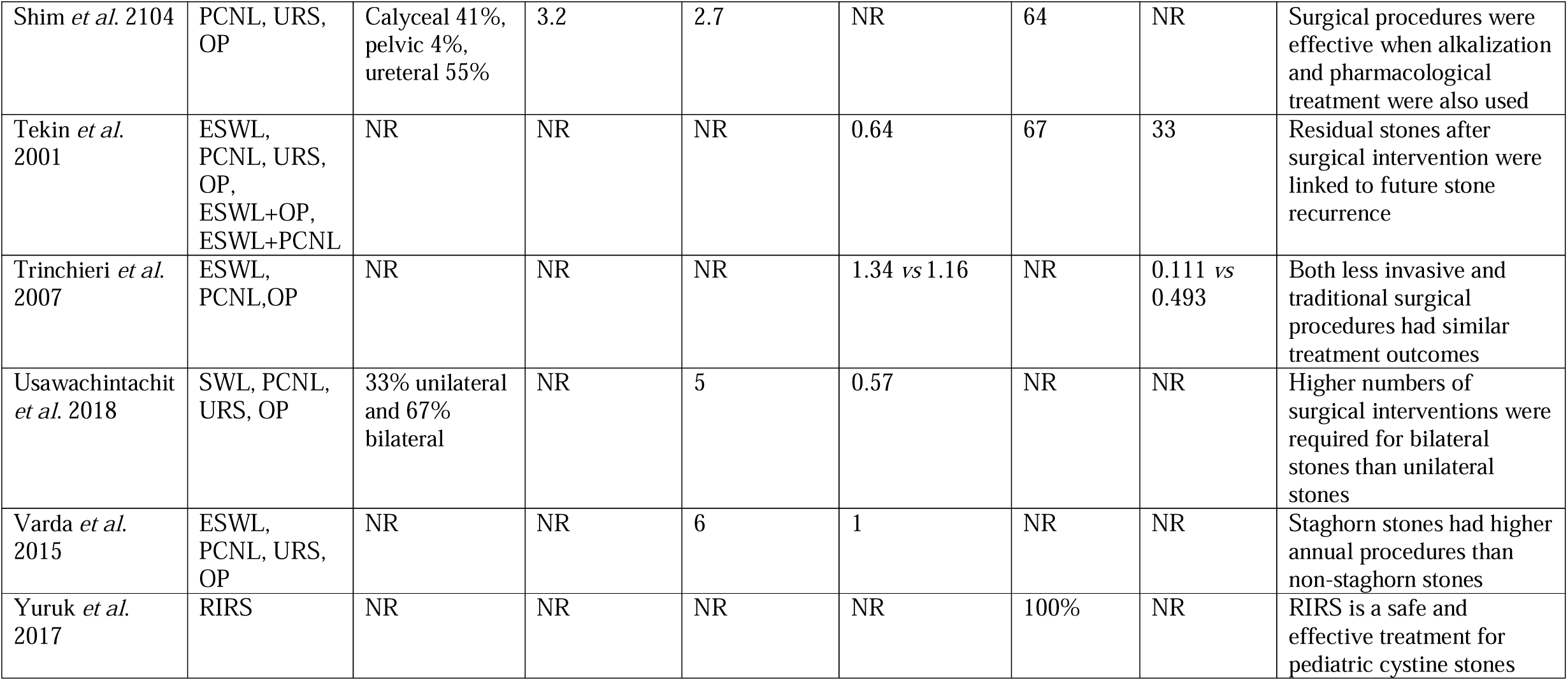
Effectiveness of surgical interventions.

| Study | Interventions | Stone position | Stone events per patient | Surgical procedures per patient | Surgical procedures per patients-year | Stone-free rate (%) | Stone recurrence rate (%) | Key outcomes |
| --- | --- | --- | --- | --- | --- | --- | --- | --- |
| Ahmed *et al.* 2008 | ESWL, PCNL, URS, OP | NR | 4.2 | 7.9 | NR | 93 | NR | Repeated surgical procedures were required even with pharmacological intervention |
| Barbey *et al.* 2000 | SWL, PCNL, RIRS, OP | 22% unilateral and 30% bilateral | 2.4 | 1.5 | 0.14 vs 0.29, $p<0.01$ | 22 | 78 | Number of surgical procedures decreased after pharmacological intervention |
| Chow *et al.* 1996 | ESWL, PCNL, RIRS | 75% unilateral and 13% bilateral | 5.5 | 0.84 | 0.3 | 75 | NR | Stone events and interventions decreased when alkalization and thiols were used |
| Chow *et al.* 1998 | ESWL, PCNL, RIRS, PCNL+ESWL | NR | NR | 1.98 | NR | 87 | 68 | Stone free recurrence probability was higher at 1 year than 5 years after first surgical procedure |
| Moore *et al.* 2019 | SWL, PCNL, URS | NR | NR | 2.7 | NR | NR | 62.5 | Stone recurrence rate and number of procedures were lower when patients achieved stone-free status in the first treatment |
| Onal *et al.* 2013 | SWL, PCNL, URS, OP, PCNL+OP | 46% left kidney and 54% right kidney | NR | NR | NR | 63 | 31 | PCNL was a safe and effective treatment for children with cystine stones |
| Pareek *et al.* 2005 | SWL, PCNL, URS | NR | NR | 1 per patient in complaint group, $p<0.05$ | NR | 73% of compliant patients | NR | Fewer numbers of surgical interventions were required with compliant patients |
| Shim *et al.* 2104 | PCNL, URS, OP | Calyceal 41%, pelvic 4%, ureteral 55% | 3.2 | 2.7 | NR | 64 | NR | Surgical procedures were effective when alkalization and pharmacological treatment were also used |
| Tekin *et al.* 2001 | ESWL, PCNL, URS, OP, ESWL+OP, ESWL+PCNL | NR | NR | NR | 0.64 | 67 | 33 | Residual stones after surgical intervention were linked to future stone recurrence |
| Trinchieri *et al.* 2007 | ESWL, PCNL, OP | NR | NR | NR | 1.34 vs 1.16 | NR | 0.111 vs 0.493 | Both less invasive and traditional surgical procedures had similar treatment outcomes |
| Usawachintachit *et al.* 2018 | SWL, PCNL, URS, OP | 33% unilateral and 67% bilateral | NR | 5 | 0.57 | NR | NR | Higher numbers of surgical interventions were required for bilateral stones than unilateral stones |
| Varda *et al.* 2015 | ESWL, PCNL, URS, OP | NR | NR | 6 | 1 | NR | NR | Staghorn stones had higher annual procedures than non-staghorn stones |
| Yuruk *et al.* 2017 | RIRS | NR | NR | NR | NR | 100% | NR | RIRS is a safe and effective treatment for pediatric cystine stones |

Stone events were 2.4,^17^ 3.2,^32^ 4.2^10^ and 5.5^19^ per patient for the treatment period in 4 studies. Total surgical procedures ranged from 0.84 to 7.9 per patient.^10, 17, 19, 25, 29, 32, 33, 36, 39^ Further surgical procedures vaired from 0.29 to 1 per patient-year in 6 studies.^11, 17, 19, 33, 38, 39^ 67% of cystinuria patients had their stones removed after a single or combined ESWL, PCNL and OP intervention, and 33% still had residual stones even after surgery. The overall stone recurrence rate was 0.64 per patient-year.^11^ There were no differences between less-invasive and traditional surgical procedures with surgical procedures per patient-year (1.34 *vs* 1.16) and stone recurrence rate (0.111 *vs* 0.493), respectively.^38^

10 studies reported the percentage of stone-free patients after the first surgical procedures which varied from 22% to 100%.^10, 11, 17, 19, 20, 25, 30, 32, 36^ Stone-free rate was dependent on the type of intervention used; PNCL (85%) was more successful than other procedures, however, stone free status was not linked to probability of recurrence of stones.^25^ Stone free rate after first PCNL was 0% in cystine stone group and 40% in struvite stone group, and final stone free rate after a secondary procedure was increased to 70% in cystine stone group and 80% in struvite stone group.^40^ RIRS had a 100% stone free rate and it was considered a safe intervention in pediatric cystine stone management.^20^

5 studies reported pharmacological interventions and stone recurrence in cystinuria patients, which ranged from 31 to 78%.^11, 17, 25, 29, 30^ Overall stone recurrence was 41.1% during mean follow-up period of 2.5 years and recurrence rate was decreased to 16.6% for medical prophylaxis, although, 100% recurrence for non-prophylaxis cystinuria patients.^26^ Stone recurrence was 3.2 per patient during the 5 years of follow-up and number of surgical intervention was 2.7 per patient.^32^ The recurrence rate after achieving stone-free status with surgical treatment was 31.2%, and the regrowth rate for children with residual stones was 29.4%.^30^ Moore *et al*., reported 62.5% of cystinuria patients had recurring stones during the mean follow-up period of 8.6 years and lower stone recurrence rate in patients who had stone-free status following their first treatment compared to stone-free status following multiple sessions (*p*=0.091).^29^

### Comparison between cystine and non-cystine stone patients

4 studies reported comparisons between cystine and non-cystine patients (Table 7).^34, 35, 37, 40^ Cystine stones occurred in early age compared to calcium oxalate (CaOx) stones (15.0±11.1years *vs* 33.5±14.8years, *p*<0.001), and were larger in size compared with CaOx stones (24.6±11.0mm *vs* 15.2±8.25mm, *p*<0.001).^37^ 2 studies reported cystine stone patient required higher surgical procedures than CaOx stone patients (14.1% *vs* 2.96%, *p*=0.007^35^ and 9.80±9.17 *vs* 2.73±1.95, *p*<0.001^37^). Cystine stone patients underwent significantly more surgical procedures than non-cystine stone patients (4.0 *vs* 1.86, *p*<0.001).^34^ Similar findings were reported by Yamacake *et al*., where cystine stone patients required higher surgical procedures than struvite stone patients (3.57±1.04 *vs* 2.0±1.22, *p*=0.028).^40^

**Table 7.** Comparison between cystine and non-cystine stone patients.

| Study | Interventions | Comparison | No. of surgical procedures (cystine vs non-cystine stones) | Serum creatinine (cystine vs non-cystine stones) | Stone size (mm) (cystine vs non-cystine stones) | Age (years) (cystine vs non-cystine stones) | Key outcomes |
| --- | --- | --- | --- | --- | --- | --- | --- |
| Assimos *et al.* 2002 | ESWL, PCNL, OP | Cystine and CaOx stone patients | 14.1% vs 2.96%, $p=0.007$ | Higher serum creatinine levels in cystinuria patients, $P=0.0001$ | NR | $P=0.00001$ | Higher numbers of surgical procedures were required to remove cystine stones than CaOx stones |
| Sfoungaristos *et al.* 2015 | ESWL, PCNL, URS, RIRS | Cystine and CaOx stone patients | 9.80±9.17 vs 2.73±1.95, $p<0.001$ | 110.97±50.4mmol/L vs 80.7±18.9mmol/L, $p<0.005$ | Larger in size, 24.6±11.0mm vs 15.2±8.2mm, $p<0.001$ | Early age, 15.0±11.1 vs 33.5±14.8, $p<0.001$ | Cystine stones were larger than CaOx stones in size and present at an earlier age |
| Worcester *et al.* 2006 | Potassium citrate<br>D-penicillamine or Captopril, Tiopronin<br>SWL | Cystine and non-cystine stone patients | Before therapy (4.0 vs 1.86, $p<0.001$) and after therapy (0.88 vs 0.23, $p<0.001$) | NR | NR | NR | A higher number of surgical procedures were required for treatment of cystine stones compared to non-cystine stones |
| Yamacake *et al.* 2016 | PCNL, SWL and OP | Cystine and struvite stone patients | 3.57±1.04 and 2.0±1.22, $p=0.028$ | 1.05±0.65mg/dL and 0.98±0.53mg/dL, $p=0.601$ | NR | NR | A higher number of stone-free procedures were required for cystine stone patients than struvite stone patients |

2 SWL/ESWL, extracorporeal shockwave lithotripsy; PCNL, percutaneous nephrolithotomy; RIRS, retrograde intrarenal surgery; URS, 3 ureterorenoscopy; OP, open surgery; CaOx, calcium oxalate; NR, not reported; NS, not significant

The average number of surgical procedures in cystinuria patients was 3.1 (range 1-8/patient).^29^ Thiol intervention decreased the number of required surgical procedures from 0.88 to 0.23 (*p*<0.001) during the follow-up period.^34^ Furthermore, SWL (1.18±1.56 and 0.73±1.02, *p*=0.0001), PCNL (0.93±1.18 and 0.09±0.47, *p*=0.00001) and OP (0.92±1.21 and 0.16±0.55, *p*=0.0001) procedures were reported for cystine stone than CaOx stone patients.^35^ Similar findings were reported by Sfoungaristos *et al*., with cystinuria patients more frequently receiving SWL (6.33±6.63 *vs* 1.63±1.73, *p*<0.001) and PCNL (1.53±1.41 *vs* 0.37±0.49, *p*<0.001) than CaOx stone patients.^37^ Overall the number of surgical procedures to remove stones was higher in cystine stone patients than CaOx stone patients (9.80±9.17 *vs* 2.73±1.95, *p*<0.001).^37^

### Patient compliance and treatment outcomes

The management and outcomes of cystinuria patients depends on patient compliance. 3 studies reported patient compliance with follow-up protocols based on their diet, medication regimen and frequency of patient visits to the clinic/doctor following intervention.^32, 36, 37^ Medical compliance after the diagnosis of cystine stones was 64.3% and overall success of the treatment was 64.2%.^32^ A monthly, quarterly and semi-annual clinic visit was required for medical evaluation of cystinuria patients, and stone-free rates were higher (73%) in patients that were compliant to treatment compared to non-compliant patients (33%).^36^ The average number of surgical interventions was 1.0 in compliant patients compared to 4.0 in non-compliant patients (*p*<0.05).^36^ The average serum creatinine (1.02mg/dL *vs* 1.1mg/dL, *p*=0.32) and urinary cystine excretion (282.2±52.6mg/L *vs* 382.4±61.3mg/L, *p*=0.25), and urinary pH (7.4±0.2 *vs* 7.1±0.3, *p*=0.66) were different for compliant and non-compliant cystinuria patients.^36^ There were lower compliance rates to diet (30 *vs* 62.5%) and medical treatment (64.3 *vs* 85.7%) for cystine stone patients compared with CaOx stone patients.^37^ Cystinuria patient compliance was associated with side effects of pharmacological agents and surgical procedures.^37^

## DISCUSSION

In this systematic review, we synthesized the results from the observational studies of existing intervention approaches for cystinuria including hydration and diuresis, alkalization, pharmacological and surgical procedures. The initial management of cystinuria patients includes hydration and urine alkalization to decrease urinary cystine levels to <250-300mg/L. Fluid intake should maintain urine volume >3L/day. Potassium citrate and sodium bicarbonate can then be used to increase urine pH >6.5, and cystine excretion to <250mg/L.^11, 18^ Additional pharmacological interventions are introduced if cystinuria patients do not respond to hydration and alkalization therapy. These additional interventions typically include treatment with tiopronin, D-penicillamine and captopril.^19^ Pharmacological treatments are effective in lowering urinary cystine levels ultimately decreasing stone events, stone recurrence rate, and increasing stone free rates in cystinuria patients.^11, 17, 19, 23-25, 27-29, 31^ While alkalization and pharmacological interventions are somewhat effective at decreasing stone episodes, these interventions are often poor at reducing cystine stone recurrence. Therefore, cystinuria patients often require surgical interventions to decrease stone recurrence rates and stone growth. To maintain a stone-free status, combined approaches of thiol-drugs and surgical procedures are required.^10, 11^ Multiple studies report that cystinuria patients require significantly higher numbers of surgical interventions than other urinary stone patients.^34, 35, 37, 40^ A greater number of ESWL, PCNL and OP procedures were required for cystine stones than CaOx stones and patients were at increased risk for renal impairment and loss of renal function.^35, 37^ PCNL was the most successful surgical procedure for achieving stone-free status in cystinuria patients.^25^ PCNL is also a safe and effective treatment for children with cystine stones.^30^ Although existing interventions are useful for reducing cystine stones, patient compliance with pharmacological medications is poor due to adverse side effects and high recurrence rates of cystine stones. Hence, management of cystine stones requires improved intervention approaches to reduce stone recurrence rates.^38^

Current treatment strategies for cystinuria are focused on relieving symptoms and preventing the formation of cystine stones. It is accepted that increasing fluid intake, regular monitoring of urinary pH and reduced dietary methionine and salt intake can reduce urinary cystine concentration.^41^ This can be achieved by high fluid consumption that increases urine volume to wash out cystine crystals, and alkalinizing supplements that maintain urine pH >6.5 to inhibit cystine aggregation and reduce cystine stone development.^18^ Usually, cystine is poorly soluble between pH 5-6.5 and cystine stones formed when individual cystine excretion in urine is exceeding 240-300mg/L (1.33-1.66mmol/L).^1^ Therefore, urine pH of >6.5 may help the dissolution of stones.^18^ Hence, urinary pH must be routinely monitored. Hydration and alkalization therapies prescribed according to body weight and severity of disease may improve quality of life of cystinuria patients by controlling urinary cystine concentration.

Thiol-based drugs are capable of breaking the disulfide bond of cystine producing cysteine, which is more soluble in urine and inhibits stone formation.^1^ These thiol drugs combine with cystine to form a soluble disulfide complex that prevents stone formation and promotes stone dissolution. Tiopronin, D-penicillamine and captopril are the most commonly reported thiol drugs for cystine stone management. Tiopronin has been shown to have high efficacy and less side effects compared with D-penicillamine and captopril.^42, 43^ D-penicillamine is equally effective, although tiopronin has a lower incidence of side effects compared to D-penicillamine. Typically, 75-100mg captopril is effective in patients who are non-responsive to standard treatment and in associated hypertensive cystinuria patients, but does increase the risk of worsening chronic kidney disease.^41^ Approximately 50% of cystinuria patients experience adverse effects to these medications including gastrointestinal intolerance, nephrotic syndrome, rash and leukopenia, which impacts on patient compliance to these intervetions.^41^

Surgical interventions for cystine stones are the same as those considered for other urinary stones,^41^ although those experiencing cystine stones are more likely to undergo surgical interventions more often due to the higher rate of reoccurrence.^35, 40^ The most common surgical interventions that are recommended for cystinuria include ESWL, PCNL, URS, RIRS and OP. ESWL is an effective treatment for cysteine stones smaller than 2.5 cm whereas PCNL is most useful for larger stones.^44^ During the surgical interventions, ESWL increased stone free rates in cystinuria patients, however, it is still lower than non-cystine stone patinets.^34^ Due to the composition of cystine stones and the amino acid structure, most of patient require multiple ESWL to dissolve of the stones. ESWL is effective and less invasive treatment of cystinuria preventing long-term complications of recurrent cystine stones.^45^ On the other hand, PCNL is considered a better surgical treatment of larger cystine stones where ESWL is less effective. However, PCNL is improved with ureteroscopic design and can be considered for larger and more complex stones. To achieve high stone-free rates, the majority of stones can be treated by ESWL, PCNL, URS, RIRS or OP alone or in combination.^11, 44^ URS is minimally invasive procedure usually done when ureter is examined for stone identification and this procedure is effective to observe cystine stone during blockage or stone pass out through urinary tract.^39^ URS is very safe and effective for recurrent cystine stone to perform precise surgical intervention to stone clearance.^46^ Most studies suggest RIRS is the most commonly used procedure and is a safe operative intervention. RIRS is effective, minimally invasive, safe treatment procedure in pediatric with 100% stone free rate.^20^ Usually, RIRS is used as an initial procedure more often in children, whereas ESWL and PCNL are more common procedures in adults.^47^ OP is restricted to when ESWL, PCNL, URS, RIRS alone or combined are not effective. OP procedures are recommended for complex staghorn and multiple stones, where other treatment strategies fail. PCNL and RIRS are safe and effective treatment procedures for pediatric cystine stone patients.^20, 30^

This review has several limitations, including the heterogeneity among selected studies including sample size and the short follow up time of the selected studies. Typically, RCTs are selected for systematic reviews because of their validity and strong outcomes. In this review, half of the available literature are retrospective studies without a control group. Due to the lack of RCTs studies in cystinuria, owing to the condition being rare, this review included all interventional studies having meaningful outcomes. Serum cystine levels and urinary cystine levels were not available for all included studies. Indeed, further RCTs are required for assessing the effectiveness of interventions in cystinuria.

## CONCLUSIONS

This systematic review highlights that cystinuria is a challenging disease to treat, requiring a range and/or combination of intervention strategies including hydration, alkalization, pharmacological and surgical procedures. Cystinuria patients have a high rate of cystine stone recurrence. New emerging therapies based on a more detailed understanding of cystinuria pathogenesis and treatment should focus to reducing recurrent stones, decreasing side effects, and preservation of renal function. The currently available observational studies do not have enough interventional details and long-term (>1 year) follow up, and most contain retrospective data without control groups. RCTs with long-term follow up and large sample size would be the appropriate design to obtain clear outcomes for optimal treatment strategies for cystinuria. The future successful management of cystinuria patients will require improved strategies for patient compliance and better treatment outcomes.

## Data Availability

All data used in this systematic review is available online through relevant cited references

## REFERENCES

1. Knoll T, Zöllner, A, Wendt-Nordahl G et al: Cystinuria in childhood and adolescence: recommendations for diagnosis, treatment, and follow-up. Pediatric nephrology (Berlin, Germany) 2005; 20:19.

2. Pereira DJ, Schoolwerth A C, Pais VM: Cystinuria: current concepts and future directions. Clin Nephrol 2015; 83:138.

3. Weinberger A, Sperling O, Rabinovitz M et al: High frequency of cystinuria among Jews of Libyan origin. Hum Hered 1974; 24:568.

4. Chillaron J, Font-Llitjos M, Fort J et al: Pathophysiology and treatment of cystinuria. Nat Rev Nephrol 2010; 6:424.

5. Andreassen KH, Pedersen KV, Osther SS et al: How should patients with cystine stone disease be evaluated and treated in the twenty-first century? Urolithiasis 2016; 44:65.

6. Milliner DS, Murphy ME: Urolithiasis in pediatric patients. Mayo Clin Proc 1993; 68:241.

7. Mandel NS, Mandel GS: Urinary Tract Stone Disease in the United States Veteran Population. II. Geographical Analysis of Variations in Composition. Journal of Urology 1989; 142:1516.

8. Leusmann DB, Blaschke R, Schmandt W: Results of 5035 Stone Analyses: A Contribution to Epidemiology of Urinary Stone Disease. Scandinavian Journal of Urology and Nephrology 1990; 24:205.

9. Thomas K, Wong K, Withington J et al: Cystinuria-a urologist’s perspective. Nat Rev Urol 2014; 11:270.

10. Ahmed K, Khan MS, Thomas K et al: Management of cystinuric patients: an observational, retrospective, single-centre analysis. Urologia Internationalis 2008; 80:141.

11. Tekin A, Tekgul S, Atsu N et al: Cystine calculi in children: The results of a metabolic evaluation and response to medical therapy. J Urol 2001; 165:2328–2330.

12. Calonge MJ, Gasparini P, Chillarón J et al: Cystinuria caused by mutations in rBAT, a gene involved in the transport of cystine. Nature Genetics 1994; 6:420.

13. Dello SL, Pras E, Pontesilli C et al: Comparison between SLC3A1 and SLC7A9 cystinuria patients and carriers: a need for a new classification. J Am Soc Nephrol 2002; 13:2547.

14. Feliubadaló L, Font M, Purroy J et al: Non-type I cystinuria caused by mutations in SLC7A9, encoding a subunit (bo,+AT) of rBAT. Nature Genetics 1999; 23:52.

15. Wong KA, Mein R, Wass M et al: The genetic diversity of cystinuria in a UK population of patients. BJU Int 2015; 116:109

16. Shen L, Cong X, Zhang X et al: Clinical and genetic characterization of Chinese pediatric cystine stone patients. J Pediatr Urol 2017; 13:629.e1-629e5.

17. Barbey F, Joly D, Rieu P et al: Medical treatment of cystinuria: Critical reappraisal of long-term results. Journal of Urology 2000; 163:1419.

18. Fjellstedt E, Denneberg T, Jeppsson J-O et al: A comparison of the effects of potassium citrate and sodium bicarbonate in the alkalinization of urine in homozygous cystinuria. Urological research 2001; 29:295.

19. Chow GK, Streem SB: Medical treatment of cystinuria: results of contemporary clinical practice. J Urol 1996; 156:1576–1578.

20. Yuruk E, Tuken M, Gonultas S et al: Retrograde intrarenal surgery in the management of pediatric cystine stones. Journal of pediatric urology 2017; 13:487.e1-487.e5.

21. Moher D, Shamseer L, Clarke M et al: Preferred reporting items for systematic review and meta-analysis protocols (PRISMA-P) 2015 statement. Syst Rev 2015; 4:1.

22. Slim K, Nini E, Forestier D et al: Methodological index for non-randomized studies (minors): development and validation of a new instrument. ANZ J Surg 2003; 73:712.

23. DeBerardinis RJ, Coughlin CR, Kaplan, P: Penicillamine therapy for pediatric cystinuria: experience from a cohort of American children. Journal of Urology 2008; 180:2620.

24. Dello SL, Laurenzi C, Legato A et al: Cystinuria in children and young adults: success of monitoring free-cystine urine levels. Pediatric Nephrology 2007; 22:1869.

25. Chow GK, Streem SB: Contemporary urological intervention for cystinuric implications patients: immediate and long-term impact and implications. J Urol 1998; 160:341–345.

26. Izol V, Aridogan IA, Karsli O et al: The effect of prophylactic treatment with Shohl’s solution in children with cystinuria. Journal of pediatric urology 2013; 9:1218.

27. Koichiro A, Kenichi E, Takeshi U et al: The long-term outcome of cystinuria in Japan. Urol Int 1998; 61:86–89.

28. Malieckal DA, Modersitzki F, Mara K et al: Effect of increasing doses of cystine-binding thiol drugs on cystine capacity in patients with cystinuria. Urolithiasis 2019; 13:13.

29. Moore SL, Somani BK, Cook P: Journey of a cystinuric patient with a long-term follow-up from a medical stone clinic: necessity to be SaFER (stone and fragments entirely removed). Urolithiasis 2019; 47:165.

30. Onal B, Dogan C, Citgez S et al: Percutaneous nephrolithotomy in children with cystine stone: long-term outcomes from a single institution. J Urol 2013; 190:234.

31. Pietrow P, Auge BK, Weizer AZ et al: Durability of the medical management of cystinuria. J Urol 2003; 169:68.

32. Shim M, Park HK: Multimodal treatments of cystine stones: an observational, retrospective single-center analysis of 14 cases. Korean Journal of Urology 2014; 55:515.

33. Usawachintachit M, Sherer B, Hudnall M et al: Clinical outcomes for cystinuria patients with unilateral versus bilateral cystine stone disease. J Endourol 2018; 32:148.

34. Worcester EM, Coe FL, Evan AP et al: Reduced renal function and benefits of treatment in cystinuria vs other forms of nephrolithiasis. BJU International 2006; 97:1285.

35. Assimos DG, Leslie SW, Christopher N et al: The impact of cystinuria on renal function. The Journal of urology 2002; 168:27.

36. Pareek G, Steele TH, Nakada SY: Urological intervention in patients with cystinuria is decreased with medical compliance. J Urol 2005; 174:2250.

37. Sfoungaristos S, Hakim R, Katz R et al: Cystine stones: A single tertiary center experience. Journal of Endourology 2015; 29:357.

38. Trinchieri A, Montanari E, Zanetti G et al: The impact of new technology in the treatment of cystine stones. Urological Research 2007; 35:129.

39. Varda BK, Johnson EK, Johnson KL et al: Imaging and surgical utilization for pediatric cystinuria patients: A single-institution cohort study. Journal of pediatric urology 2016; 12:106.e1.

40. Yamacake KGR, Marchini GS, Reis S et al: The challenge of cystine and struvite stone formers: clinical, metabolic and surgical assessment. International Braz J Urol 2016; 42:977.

41. Biyani CS, Cartledge JJ: Cystinuria—Diagnosis and Management. EAU-EBU Update Series 2006; 4:175.

42. Rhodes HL, Yarram-Smith L, Rice SJ et al: Clinical and genetic analysis of patients with cystinuria in the United Kingdom. Clin J Am Soc Nephrol 2015; 10:1235.

43. Daudon M, Cohen-Solal F, Barbey F et al: Cystine crystal volume determination: a useful tool in the management of cystinuric patients. Urological Research 2003; 31:207.

44. Cranidis AI, Karayannis AA, Delakas DS et al: Cystine stones: The efficacy of percutaneous and shock wave lithotripsy. Urologia Internationalis 1996; 56:180.

45. Katz G, Kovalski N, Landau EH: Extracorporeal shock wave lithotripsy for treatment of ureterolithiasis in patients with cystinuria. British journal of urology 1993; 72:13.

46. Ruggera L, Zanin M, Beltrami P et al: Retrograde transureteral approach: a safe and efficient treatment for recurrent cystine renal stones. Urological Research 2011; 39:411.

47. Azili MN, Ozcan F, Tiryaki T: Retrograde intrarenal surgery for the treatment of renal stones in children: Factors influencing stone clearance and complications. Journal of Pediatric Surgery 2014; 49:1161.

